# Red Blood Cell-Conditioned Media from Non-Alcoholic Fatty Liver Disease Patients Contain Increased MCP1 and Induce TNF-α Release

**DOI:** 10.1101/2021.05.10.21256939

**Authors:** Charalampos Papadopoulos, Konstantinos Mimidis, Dimitris Papazoglou, George Kolios, Ioannis Tentes, Konstantinos Anagnostopoulos

## Abstract

**BACKGROUND:** Non-Alcoholic Fatty Liver Disease (NAFLD) constitutes a global pandemic. An intricate network among cytokines and lipids possesses a central role in NAFLD pathogenesis. Red blood cells comprise an important source of both cytokines and signaling lipids and have an important role in the molecular crosstalk during immunometabolic deregulation. However, their role in NAFLD has not been investigated in deep.

**METHODS:** Conditioned media from erythrocytes derived from 10 NAFLD patients (4 men, 6 women, aged 57.875±15,16) and 10 healthy controls (4 men, 6 women, aged 39.3±15.55) were produced and used for the analysis of 9 cytokines (IFN-γ, TNF-α, CCL2, CCL5, IL-8, IL-1β, IL-12p40, IL-17, MIP-1β), 2 signaling lipids (Sphingosine 1-phosphate and Lysophosphatidic Acid), and cholesterol. Their effect on the cytokine profile released by RAW 264.7 macrophages was also studied.

**RESULTS:** Erythrocytes from patients with NAFLD augmented the levels of MCP1 in the growth medium in comparison to the erythrocytes derived from healthy controls (37±40 pg/ml vs 6.51±5.63). No statistically significant differences were found between patients and healthy controls with regard to S1P, LPA, cholesterol and 8 other cytokines. TNF-a release by RAW 264.7 cells was increased after incubation with patient-derived erythrocyte conditioned medium compared to medium without RAW 264.7 cells from either healthy of NAFLD subjects.

**CONCLUSIONS:** Erythrocytes could contribute to the liver infiltration by monocytes and to the activation of macrophages, partially due to release of CCL2, in the context of NAFLD.

## 1. Introduction

Non-alcoholic fatty liver disease (NAFLD) is defined as accumulation of fat in at least 5% of hepatocytes, after the exclusion of other secondary causes, such as significant alcohol consumption, use of steatogenic medication and hereditary, autoimmune or viral hepatic disorders. NAFLD is histologically further categorized into nonalcoholic fatty liver - NAFL (presence of hepatic steatosis with no evidence of hepatocellular injury in the form of ballooning of the hepatocytes) and nonalcoholic steatohepatitis – NASH (presence of hepatic steatosis and inflammation with hepatocyte injury (ballooning) with or without fibrosis) (1). NASH can evolve to fibrosis, cirrhosis and even hepatocellular carcinoma (2). However, the most frequent cause of mortality of NAFLD patients is the cardiovascular complications (3).

NAFLD pathogenesis is orchestrated by an interplay of diverse molecular mechanisms. The multiparallel hits hypothesis proposed more recently, suggests that NASH is the result of numerous conditions acting in parallel, including genetic predisposition, abnormal lipid metabolism and trafficking, oxidative stress, lipotoxicity, mitochondrial dysfunction, altered production of cytokines and adipokines, gut dysbiosis and endoplasmic reticulum stress (4).

An intricate network among cytokines and lysophospholipids plays a central role in NAFLD pathogenesis. Deregulation of the synthesis and signaling of various cytokines and chemokines, such as TNF-a, CCL2, CCL5 and IL-8, has been associated with the development and progression of NAFLD (5). In addition, Sphingosine 1 Phosphate (S1P) is increased in experimental NASH animal models (6), and its binding to its receptor S1PR1, results in the activation of NFKβ and the expression of pro-inflammatory cytokines, such as tumor necrosis factor a (TNF-α) and monocyte chemoattractant protein 1 (MCP1) (7). Furthermore, the role of S1P in steatohepatitis is highlighted by the fact that administration of S1P antagonists to experimental NASH animals eradicated the disease (8). Another signaling lipid, lysophosphatidic acid (LPA), also contributes to NAFLD pathogenesis: serum LPA levels were found to be associated with the degree of hepatic fibrosis and steatosis (8,9). Finally, hepatic cholesterol accumulation contributes to mitochondrial dysfunction, endoplasmic reticulum stress, inflammation and fibrosis (10).

Apart from their typical role as oxygen transporters, human erythrocytes retain a considerable immunomodulatory capacity (11,12). Interestingly, erythrocytes constitute important pools of cytokines, chemokines, S1P, LPA and Cholesterol in the blood (13).

Despite the fact that erythrocyte dysfunction is implicated in the molecular mechanisms of NAFLD (14), not much work has been done in order to study in depth the molecular phenomena for the involvement of the erythrocytes in the pathogenesis of NAFLD. It is known that during hepatic steatosis there is a remarkable erythrocyte accumulation in the liver both *ex vivo* and *in vivo* (15). These erythrocytes externalize their phosphatidylserine (PSer) as a response to oxidative stress, which is then recognized by Kupffer cells’ receptor, lactadherin, resulting in erythrophagocytosis. This interaction leads to an increase of hepatic oxidative stress and escalates inflammation (15). In addition, Unruh et al (16) have shown that high-fat diet increases erythrocyte membrane cholesterol, externalized phosphatidylserine, bound MCP1, and ROS. These erythrocytes also induce a proinflammatory phenotype in macrophages from both control and high-fat diet-fed animals, and monocyte adhesion to the endothelium.

Taken together, these observations indicate that the erythrocyte may act as a mediator of immunometabolic interactions in the context of metabolic disease. Hence, in this study, we sought to investigate whether erythrocytes could participate in NAFLD pathogenesis by further, yet unexplored, immunomodulatory interactions with macrophages, through the release of cytokines, S1P, LPA and cholesterol.

## 2. Materials and Methods

### 2.1 Materials

Dulbecco’s Phosphate Buffered Saline (PBS) w/o Calcium w/o Magnesium - 500ml and RPMI 1640 were purchased from Biosera, France. MILLIPLEX MAP Human Cytokine/Chemokine Magnetic Bead Panel, NINE PLEX was purchased from Millipore, USA. VPC23019 and AM966 were purchased from Cayman Chemicals, USA. Human Lysophosphatidic acid ELISA kit was purchased from Cubasio, USA, and general sphingosine 1 phosphate ELISA kit was purchased from Fine biotech, China. Cholesterol colorimetric assay was from Reagent Genie, Ireland.

### 2.2 Patients

10 patients (4 men, 6 women, age 57.86±15.17) and 10 healthy controls (4 men, 6 women, age 39.3±15.55) participated in our study. They were recruited by the 1st Pathology Clinic, Department of Medicine, Democritus University of Thrace, Alexandroupolis, Greece. All patients presented hepatic steatosis according to ultrasonography. After exclusion of viral, alcoholic, drug and other causes, patients were evaluated by non-invasive biomarkers (NFS, FIB4, AST/ALT ratio). Their anthropometric characteristics are shown in Table 1. Our study was approved by the scientific council of the University Hospital of Alexandoupolis and the Ethics Committee after the informed consent of the participants.

**Table 1.**
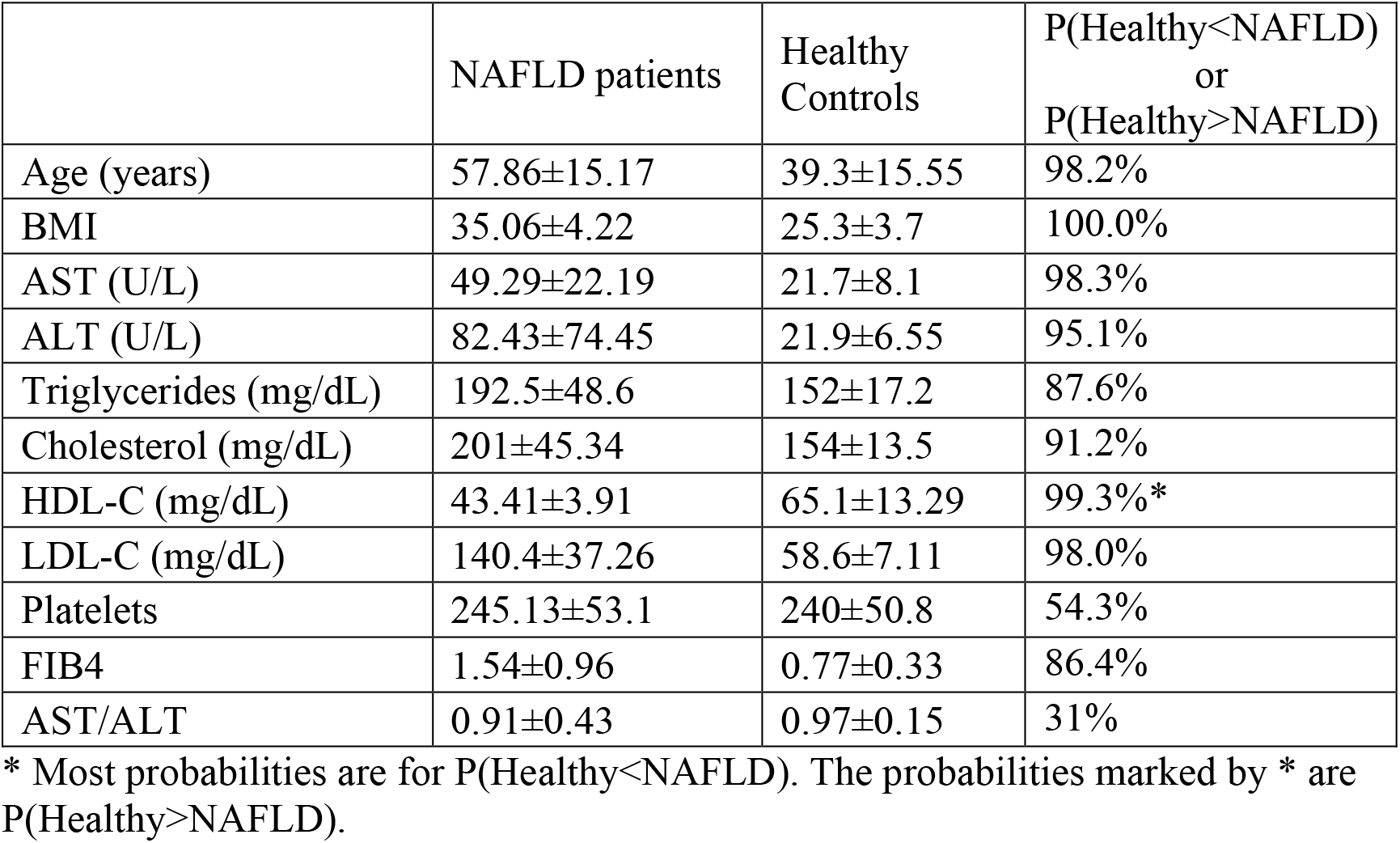
Basic characteristics of NAFLD patients and healthy subjects. P(Healthy<NAFLD) or P(Healthy>NAFLD) is the posterior probability that the difference between means of healthy subjects and NAFLD patients if less or greater than zero, respectively. (ALT: alanine aminotransferase; AST: aspartate aminotransferase; FIB4: fibrosis 4; HDL-C: high density lipoprotein cholesterol; LDL-C: low density lipoprotein cholesterol, FIB4: Non-Alcoholic Fatty Liver Disease Fibrosis score)

|  | NAFLD patients | Healthy Controls | P(Healthy<NAFLD)<br>or<br>P(Healthy>NAFLD) |
| --- | --- | --- | --- |
| Age (years) | 57.86±15.17 | 39.3±15.55 | 98.2% |
| BMI | 35.06±4.22 | 25.3±3.7 | 100.0% |
| AST (U/L) | 49.29±22.19 | 21.7±8.1 | 98.3% |
| ALT (U/L) | 82.43±74.45 | 21.9±6.55 | 95.1% |
| Triglycerides (mg/dL) | 192.5±48.6 | 152±17.2 | 87.6% |
| Cholesterol (mg/dL) | 201±45.34 | 154±13.5 | 91.2% |
| HDL-C (mg/dL) | 43.41±3.91 | 65.1±13.29 | 99.3%* |
| LDL-C (mg/dL) | 140.4±37.26 | 58.6±7.11 | 98.0% |
| Platelets | 245.13±53.1 | 240±50.8 | 54.3% |
| FIB4 | 1.54±0.96 | 0.77±0.33 | 86.4% |
| AST/ALT | 0.91±0.43 | 0.97±0.15 | 31% |

\* Most probabilities are for P(Healthy<NAFLD). The probabilities marked by \* are P(Healthy>NAFLD).

### 2.3 Isolation of red blood cells

Three milliliters of blood containing EDTA was centrifuged at 200 g for 10 minutes at 4°C. Plasma and buffy coat were removed. Then, the erythrocyte pellet was washed with cold saline solution and centrifuged at 200 g for 10 minutes at 4^0^C. The buffy coat was removed. This step was repeated 3 times. Next, 100μl of erythrocytes from the bottom of the pellet were used for cell counting and production of conditioned media.

### 2.4 Production of red blood cell-conditioned medium

Erythrocytes were incubated in RPMI 1640, supplemented with 10% FBS, 1% streptomycin/penicillin, at 5% CO2, 37°C, for 24 hours. Then, the red blood cell-derived conditioned medium (RBC-CM), from both patients (P-RBC-CM) and healthy controls (H-RBC-CM) was collected with centrifugation at 200 g for 10 minutes. Conditioned media were stored at −80°C. As control, growth medium was placed in 6 well plates, in the same conditions, thereafter, following the same procedures as for RBC-CM. Spectrophotometric assay indicated that no hemoglobin was released by erythrocytes, excluding thus, hemolysis.

### 2.5 Raw 264.7 macrophage culture

RAW 264.7 macrophages were cultured in RPMI 1640, supplemented with 10% FBS, 1% streptomycin/penicillin. When passage 12 was reached, cells were seeded in 6 well plates, at a concentration of 2×10^5^ cells/well. When confluency reached approximately 80%, RAW 264.7 macrophages were exposed to red blood cell-conditioned medium, at 5% CO2, 37°C, for 24 hours. VPC 23019, an antagonist for S1P receptors1,3 and AM966, an antagonist for LPA receptor1, (CAYMAN, chemicals, USA) were used at a concentration of 10 μM and 25 mM respectively, after diluting with DMSO. DMSO, VPC23019 and AM966 had no effect on the cytokine profile released by RAW 264.7 macrophages.

### 2.6 Analysis of Cytokines, S1P and LPA

IFN-γ, TNF-α, CCL2, CCL5, IL-8, IL-1β, IL12, IL-17, MIP-1β were studied by multiplex technology, according to manufacturer’s instructions. S1P and LPA were analyzed by ELISA method, following the manufacturers’ instructions.

### 2.7 Cholesterol content determination

Cholesterol was determined by a colorimetric assay, following the manufacturers’ instructions.

### 2.8 Statistical analysis

Results in the text are expressed as mean ± standard deviation, unless otherwise stated. Since the sample size was small, we used a Bayesian approach for statistical analysis, which is much more suited to provide meaningful results for small datasets. Bayesian analysis does not assume large sample sizes and smaller datasets can be analyzed while retaining statistical power and precision (17,18).

Statistical analysis was done with the R programming language v. 3.6 (19). Testing for difference between means was performed using the BEST package (20). The results are reported as the probability P of the difference between the means of healthy subjects and NAFLD patients being less than zero, P(Healthy<NAFLD), or greater than zero, P(Healthy>NAFLD). This probability is more intuitive than the more convoluted meaning of the frequently used p value, which is the probability of observing the data, assuming the null hypothesis (no difference between means) is correct. Probabilities above 90% were considered statistically significant.

## 3. Results

Patients included in our study presented statistically significant higher body mass index and levels of serum transaminases, triglycerides, low-density lipoprotein and total cholesterol than healthy controls, as shown in Table 1. Meanwhile, healthy controls had higher high-density lipoprotein cholesterol. These results are indicative of the metabolic malfunction during NAFLD.

The boxplot in Figure 1 shows the levels of cytokines (Fig. 1A) and lipids (Fig. 1B) in the erythrocyte conditioned medium of healthy and NAFLD subjects. The levels of the respective parameters in plain growth medium are shown as baseline reference. Erythrocytes from patients with NAFLD augmented the levels of MCP1 in the growth medium in comparison to the erythrocytes derived from healthy controls (37±40 pg/ml vs 6.51± 5.63) P(Healthy<NAFLD) = 99.4%, as shown in Figure 1A. Also, IL-8 showed a marginal difference between means, with P(Healthy<NAFLD)=94.6%. In addition, some cytokines (IFN-γ, IL-1β, IL-17) were found to be below the detection limit of the kit. The rest of the cytokines measured did not show any statistically significant difference (MIP1β, RANTES, and TNF-α) or were lower than the detection limit of the kit (IL-1β, IL-12p40, IFN-γ).

**Figure 1.**
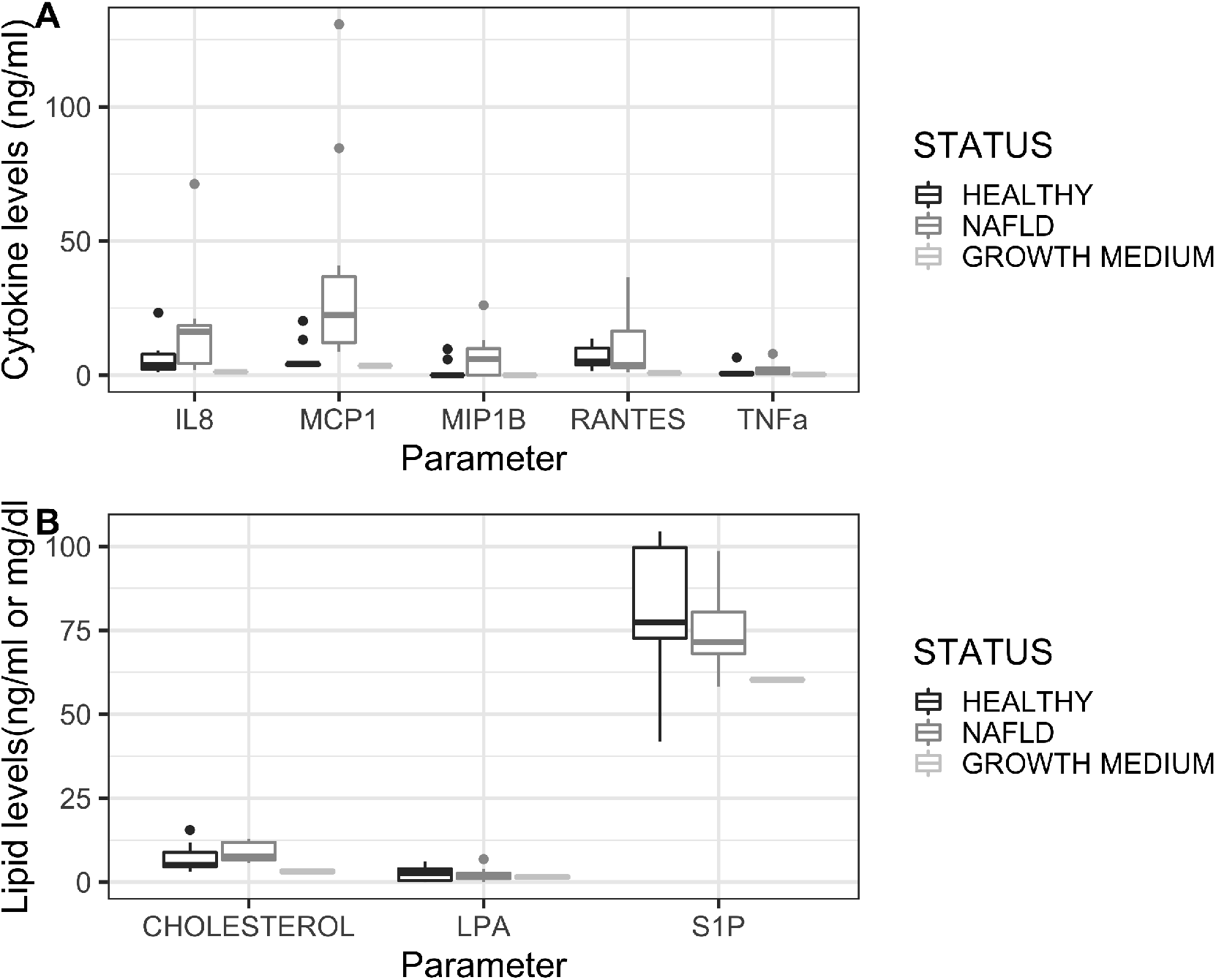
(A) Cytokine levels (ng/ml) in growth medium and red blood cell-conditioned medium (RBC-CM) from patients and healthy controls. (B): S1P and LPA (ng/ml) and cholesterol levels (mg/dl) in growth medium and red blood cell-conditioned medium from patients and healthy controls. LPA: lysophosphatidic acid; S1P: sphingosine 1 phosphate.

Since red blood cells secrete bioactive lipids, we examined the levels of S1P and LPA in RBC-CM. No statistically significant differences were found between patients and healthy controls (75.62±12.77 vs 81.18±19.72 for S1P, and 2.28±2.29 vs 2.59±2.15 pg/ml, for LPA) (Figure 1B). Similarly, cholesterol levels did not differ between the two groups (8.9±2.85 vs 4.51±5.23 mg/dl) (Figure 1B).

Next, we examined the effect of conditioned medium both from patients and healthy controls (P-RBC-CM and H-RBC-CM respectively) on the cytokine profile released by RAW 264.7 macrophages (Figure 2). TNF-α release in RAW stimulated with P-RBC-CM increased (36.84±25.21 pg/ml) in comparison to the effect of H-RBC-CM (1.67± 1.23 pg/ml), P(H-RBC-CM-RAW<P-RBC-CM-RAW) = 91.3%. This means that the patients’ erythrocyte conditioned medium induces the release of TNF-α by RAW macrophages. Also, TNF-α levels of P-RBC-CM-RAW were higher than P-RBC-CM alone (3.19±3.65 pg/ml), P(P-RBC-CM<P-RBC-CM-RAW) = 93.1%. This observation shows that the TNF-α increase is not inherent to the patients’ erythrocytes but is mainly released by the RAW macrophage cells, thus ruling-out the hypothesis that TNF-α originates from the patients’ erythrocytes.

**Figure 2.**
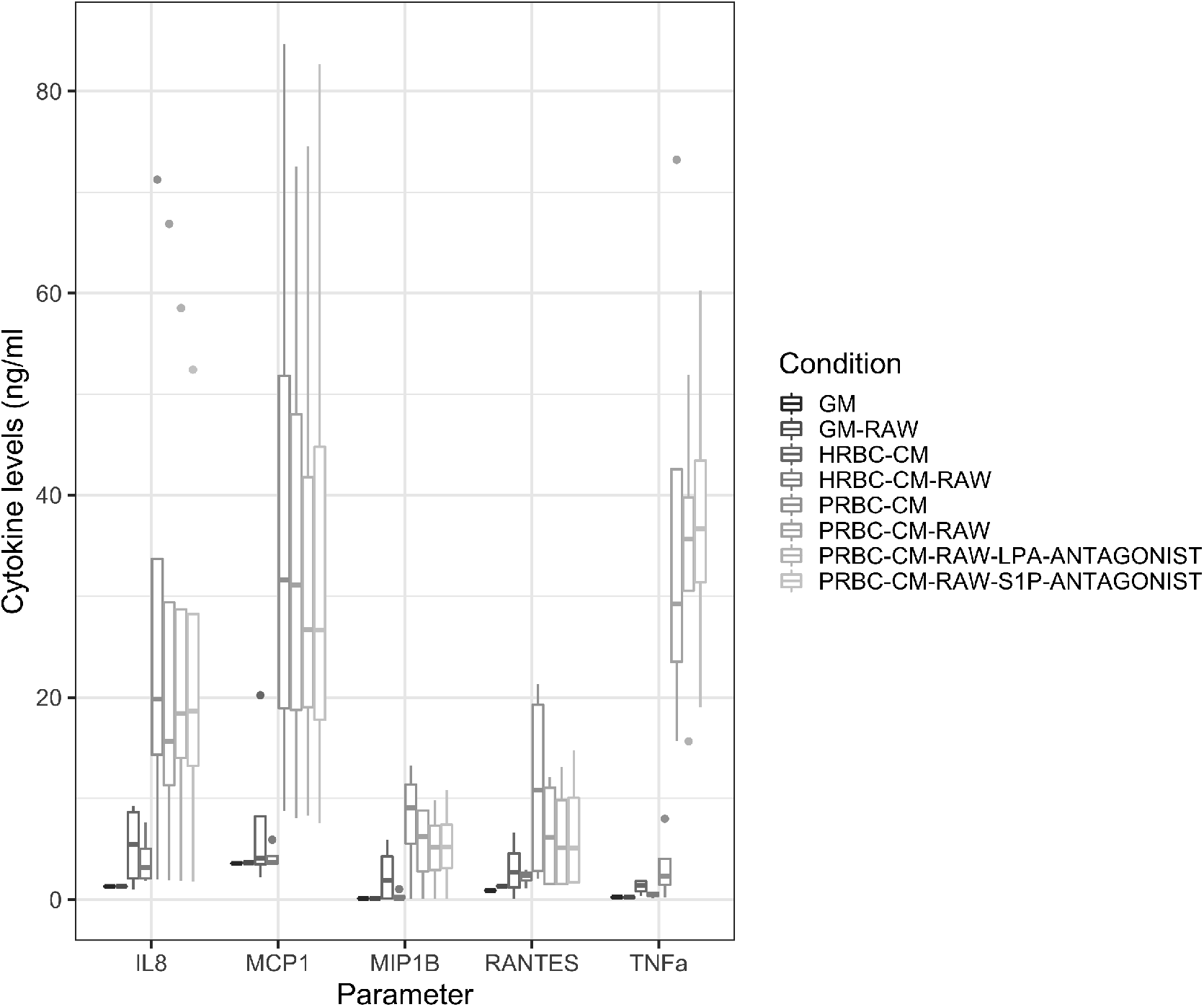
Cytokine levels released by RAW 264.7 cells after incubation for 24 hours with growth medium and red blood cell-conditioned medium from patients and healthy controls. GM: growth medium; GM-RAW; RAW 264.7 conditioned medium; H-RBC-CM: healthy control-derived red blood cell conditioned medium, H-RBC-CM-RAW: healthy control-derived red blood cell conditioned medium incubated with macrophages RAW 264.7; P-RBC-CM: patient-derived red blood cell conditioned medium; P-RBC-CM-RAW: patient-derived red blood cell conditioned medium incubated with macrophages RAW 264.7; P-RBC-CM-RAW-LPA antagonist: patient-derived red blood cell conditioned medium incubated with macrophages RAW 264.7 pre-incubated with AM966, an antagonist for LPA receptor1 at concentration of 25Mm; P-RBC-CM-RAW-S1P antagonist: patient-derived red blood cell conditioned medium incubated with macrophages RAW 264.7 pre-incubated with VPC 23019, an antagonist for S1P receptors 1 and 3, at a concentration of 10 μM.

Finally, we examined whether the effect of erythrocytes from patients with NAFLD was mediated through the action of S1P and/or LPA, using antagonists for S1P receptors 1 and 3, and LPA receptor 1 (Figure 2). There were no statistically significant differences in the profile of the secreted cytokines between cells treated with Patient-derived conditioned medium in the presence or absence of VPC23019 or AM966. Therefore, our results indicate that S1PR1, S1PR3 and LPAR1 are not implicated in the process.

## 4. Discussion

In this study, the role of red blood cells in the inflammatory response of macrophages in the context of NAFLD was investigated. We provide preliminary evidence that red blood cells possibly release chemokines and induce TNF-α release from macrophages. This effect appeared to be independent of the erythrocyte-derived S1P, LPA and cholesterol.

Darbonne et al (21), were the first to show that erythrocytes bind chemokine CXCL8 (IL-8) and CCL2 (MCP1), through the Duffy Atypical Receptor of Chemokines. Apart from chemokine scavenging, red blood cells also possess the ability to release cytokines and growth factors. Wei et al (22), showed that IL-33 is released by erythrocyte through hemolysis. Next, Karsten et al (12) showed that the erythrocytes of healthy volunteers can release more than 40 cytokines, chemokines and growth factors. In addition, in a study of Unruh et al (16), erythrocyte-bound MCP1 was found to increase as a result of high-fat diet in experimental animals, and possibly, after release, participates in monocyte chemotaxis. Based on those observations, we explored a potential chemokine and cytokine releasing property of erythrocytes in the context of NAFLD, because these signaling mediators are implicated in a substantial part of the inflammatory cellular communication. Indeed, in our study, MCP1 was found increased in NAFLD patients-derived red blood cell conditioned media. CXXL8 also displayed a small, but significant, increase.

Since MCP1 also induces pro-inflammatory cytokine expression in macrophages (23), we explored the effect of red blood cell-derived conditioned media on the cytokine profile secreted by RAW 264.7 macrophages. After examining the levels of 9 cytokines by employing multiplex technology, we found that P-RBC-CM provoked significant increase in TNF-α release. Since our protocol permitted the interaction with only erythrocyte-derived soluble factors, we hypothesized that the release of cytokines or signaling lipids could mediate the effects of red blood cells to the activation of monocytes or macrophages. In our study we excluded the role of both S1P and LPA, since their levels did not differentiate between patients and healthy controls, and receptor antagonism did not have an effect on RAW 264.7 macrophages. In addition, cholesterol levels did not differ between patients and healthy controls. However, erythrocyte-derived reactive oxygen species (ROS), free heme, hemoglobin and microvesicles have all been implicated in the regulation of monocyte and macrophage function (24–26), and cannot be excluded from our study as potent mediators of the reported TNF-α increase. In fact, erythrocyte vesiculation is increased in both metabolic syndrome and hepatic cirrhosis (27,28) and it could explain MCP1 release (29) in our study.

Apart from microvesicle release, other mechanisms explaining the augmented release of chemokines could be the differential binding capacity in the context of the disease. In particular, the levels of DARC sulfation could also lead to different capacity of binding, and thus, releasing chemokines (30). However, to what extent this applies to all chemokines remains to be elucidated.

In our results, we observe a large variance regarding the degree of MCP1 and other chemokines’ release from patients’ erythrocytes. There are numerous reasons possibly explaining this fact. Firstly, if chemokine release is mediated mainly through formation of microvesicles, then a varied release is anticipated. Xiong et al (29) showed that microvesicles formed by red blood cells present altered binding ability and a large variance with regards to the dissociation constant of chemokines. Another reason could be the high variability of erythrocytes between and within individuals. Especially, in NAFLD patients a high Red-Cell Distribution Width (RDW) is observed (31). Since RDW indicates the existence of different red blood cell subpopulations in the blood, these could have different physicochemical characteristics.

We are aware that we analyzed samples from a limited number of patients with NAFLD who were not biopsy-proven. As a result, we are missing a possible relation between NAFLD stage and cytokine profile released by erythrocytes and RAW 264.7 after interacting with RBC-CM. In view of the fact that the erythrocytes release such a plethora of signaling agents targeting a variety of cells that are implicated in the pathogenesis of NAFLD, further investigation is mandatory.

From these results, it is fair to conclude that erythrocytes can possibly contribute to liver infiltration by monocytes and the activation of macrophages partially due to release of MCP1, acting within the context of NAFLD pathogenesis. However, expansion of this study to a larger patient group and more experimental studies in animal models are needed.

## Data Availability

Data will be avaiable upon request.

## Acknowledgements

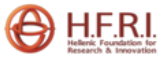

The research work was supported by the Hellenic Foundation for Research and Innovation (HFRI) under the HFRI PhD Fellowship grant (Fellowship Number:1343).

